# Serological detection of 2019-nCoV respond to the epidemic: A useful complement to nucleic acid testing

**DOI:** 10.1101/2020.03.04.20030916

**Authors:** Jin Zhang, Jianhua Liu, Na Li, Yong Liu, Rui Ye, Xiaosong Qin, Rui Zheng

## Abstract

Corona Virus Disease 2019 (COVID-19) has spread rapidly to more than 70 countries and regions overseas and over 80000 cases have been infected, resulting in more than three thousand deaths. Rapid diagnosis of patients remains a bottleneck in containing the progress of the epidemic. We used automated chemiluminescent immunoassay to detect serum IgM and IgG antibodies to 2019-nCoV of 736 subjects. COVID-19 patients were becoming reactive(positive) for specific antibodies from 7-12 days after the onset of morbidity. Specific IgM and IgG increased with the progression of the disease. The areas under the ROC curves of IgM and IgG were 0.988 and 1.000, respectively. Specific antibody detection has good sensitivity and specificity. Detection of specific antibodies in patients with fever can be a good distinction between COVID-19 and other diseases, so as to be a complement to nucleic acid diagnosis to early diagnosis of suspected cases.

## Introduction

Coronaviruses (CoVs) are enveloped single-stranded positive-sense RNA viruses, which are widely distributed in humans and other mammals. Coronaviruses usually cause respiratory, digestive and nervous system diseases in humans and animals (*1*). In the past 20 years, coronavirus has caused two global epidemics of severe respiratory infectious diseases, one of which was severe acute respiratory syndrome (SARS) (*2,3*) in Guangdong Province of China from 2002 to 2003. The other was the Middle East Respiratory Syndrome (Middle East respiratory syndrome, MERS) outbreak in Saudi Arabia in 2012 (*4*).

Pneumonia caused by 2019 novel coronavirus (2019-nCoV) was first reported in Wuhan, Hubei Province, China in December 2019. Then it spread rapidly nationwide. At the same time, more than 70 countries and regions overseas have reported for infected cases. As of 24:00 on February 29th, 2020, a total of 79824 confirmed Corona Virus Disease 2019 (COVID-19) cases and 2870 deaths have been reported in China. (http://www.nhc.gov.cn/xcs/yqfkdt/202003/9d462194284840ad96ce75eb8e4c8039.shtml).

Since the outbreak of the epidemic, the China National Health Commission has published the “Diagnosis and Treatment plan of Corona Virus Disease 2019” and has revised it several times along with the actual situation of the progress of the epidemic (*5-9*). In addition to having a history of epidemiology, clinical signs and imaging characteristics of viral pneumonia, an important diagnostic criterion for COVID-19 patients is their positive 2019-nCoV nucleic acid test result of nasal and pharyngeal swab (*5-9*). However, in the actual diagnosis and treatment, the sensitivity of nucleic acid detection was not ideal enough. Only 30-50% of the confirmed COVID-19 cases had positive results, moreover, in some confirmed case, nucleic acid testing often took four or more tests to get a positive result. It is necessary to use a fast and convenient method to realize the rapid diagnosis of 2019-nCoV infection.

After the virus infects the organism, the immune system carries on the immune defense to the virus and produces the specific antibody. In the laboratory diagnosis of infectious diseases, the detection of specific antibodies to pathogens is a sensitive method for fast diagnosis. However, how the 2019-nCoV antibody produced and changed during COVID-19 progression is still unclear.

In this study, we used automated chemiluminescent immunoassay to detect serum IgM and IgG antibodies to 2019-nCoV, to understand the process of antibody production in disease progression, and to evaluate the value of antibody detection in the laboratory diagnosis of COVID-19.

The study was conducted in accordance with the International Coordinating Council for Clinical Trials and the Helsinki Declaration, and was approved by the Hospital Ethics Review Committee (Ethics No 2020PS038K), and the patient’s informed consent was exempted.

## Method

### Study design and participants

A total of 736 subjects were included in the study. 228 suspected COVID-19 cases were admitted to the fever clinic from January 21, 2020 to February 16, 2020 in Shengjing Hospital of Chinese Medical University, which is the designated hospital for COVID-19 in Liaoning Province. All of 228 cases were observed quarantine admission and taken the nasal and pharyngeal swab for 2019-nCoV nucleic acid testing. According to the “Diagnosis and Treatment plan of Corona Virus Disease 2019” (*8*), 3 cases with positive result for 2019-nCoV nucleic acid detection were recorded as COVID-19 group. The other 225 cases that had twice negative result for the 2019-nCoV nucleic acid detection were named non-COVID-19 group.

Another 222 outpatients with other diseases in the same period, 63 medical staffs worked for fever clinic and 223 healthy physical examinees in 2018 were collected and were named other disease group, medical staff group and health control group, respectively.

#### Clinical Data Collection

According to the unified form, two residents collected clinical data from medical records separately.

#### Blood sampling

Fasting venous blood (5ml) was collected from all the subjects and put into the yellow head vacuum tube containing separation gel. After centrifugation, the serum samples were stored at-20 °C.

#### 2019-nCoV nucleic acid detection

Nasopharynx/oropharynx swab samples were collected by regional Center for Disease Control. Fluorescence Reverse transcriptase polymerase chain reaction (RT-PCR) was used to detect the expression of open reading frame 1ab (ORF1ab) and nucleocapsid protein (NCP) in 2019-nCoV genome. The CT value of 2019-nCoV nucleic acid test results should be interpreted according to the recommendations of the manufacturer’s instructions, and the suspicious results should be notified for clinical re-sampling and re-examination. In order to be diagnosed as positive in laboratory test results, it is necessary to meet the standard that 2019-nCoV ORF1ab and N gene of same sample shows at least one target specific RT-PCR test result is positive.

### Diagnostic criteria for confirmed, severe and critical cases for COVID-19

The diagnosis was made according to the “Diagnosis and Treatment plan of Corona Virus Disease 2019 (Tentative fifth revised edition)”(*8*) published by China National Health Commission.

### 2019-nCoV IgM and IgG antibody detection

Chemiluminescence detection kit from Shenzhen Yahuilong Biotechnology Co Ltd was used to detect 2019-nCoV IgM and IgG antibody. Magnetic particle coated antigens including 2019-nCoV Spike protein S and nucleocapsid protein N antigen. iFlash 3000 automatic chemiluminescence immunoassay analyzer from Shenzhen Yahuilong Biotechnology Co Ltd. All operations are carried out after strict calibration and quality control in accordance with the operator’s instructions. The results will be reported in 30 minutes after sample loading by relative luminescence intensity unit(RLU). There was a positive correlation between the amount of 2019-nCoV IgM or IgG antibody in the sample and RLU, and the concentration of 2019-nCoV IgM or IgG antibody (AU/ml) was automatically calculated according to RLU and built-in calibration curve, and 10.0 AU/ml was regarded as reactive (positive).

### Statistical analyses

SPSS version 20.0 (SPSS Inc., Chicago, IL) and GraphPad Prism version 5.01 (GraphPad Software Inc., San Diego, CA) were used for statistical analyses. Quantitative variables were expressed as median (P99). The normality of variables was tested using Kolmogorov–Smirnov test. and LDS-t-test for comparison among groups. A receiver operating characteristic curve (ROC) was plotted to evaluate the diagnostic performance and correlations determined by Spearman’s rank correlation. Z test was used to compare AUC between two groups. All tests were two sided and *P* values < 0.05 were considered statistically significant.

## Results

Of the 3 cases of confirmed case, 2 were male and 1 was female. The age ranged from 39 to 57 years. 2 patients had diabetes and hypertension respectively. 1 was a common case, 2 were severe cases. 1 case had history of Wuhan contact and the other 2 cases had no clear epidemiological history (Table 1).

**Table1.**
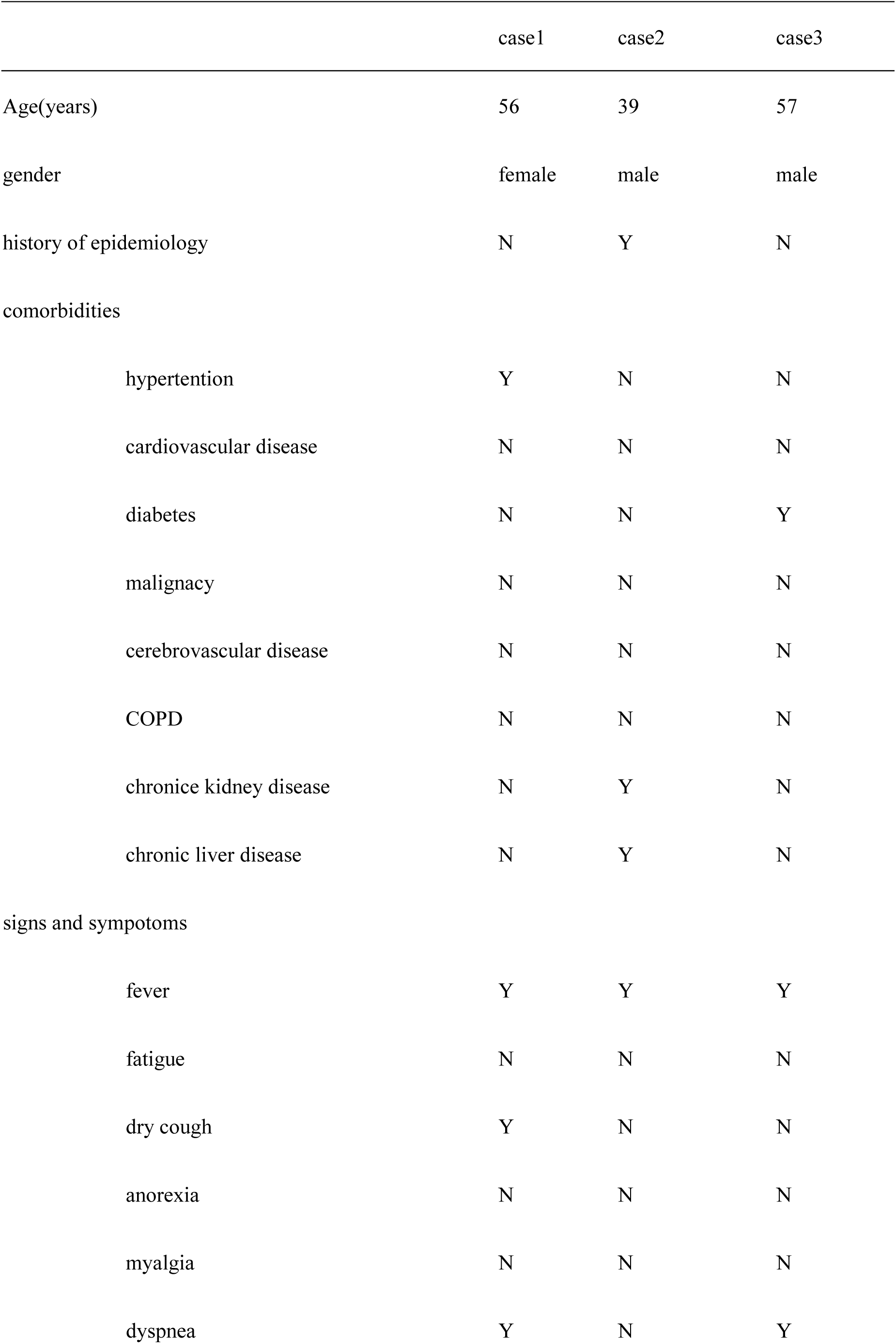
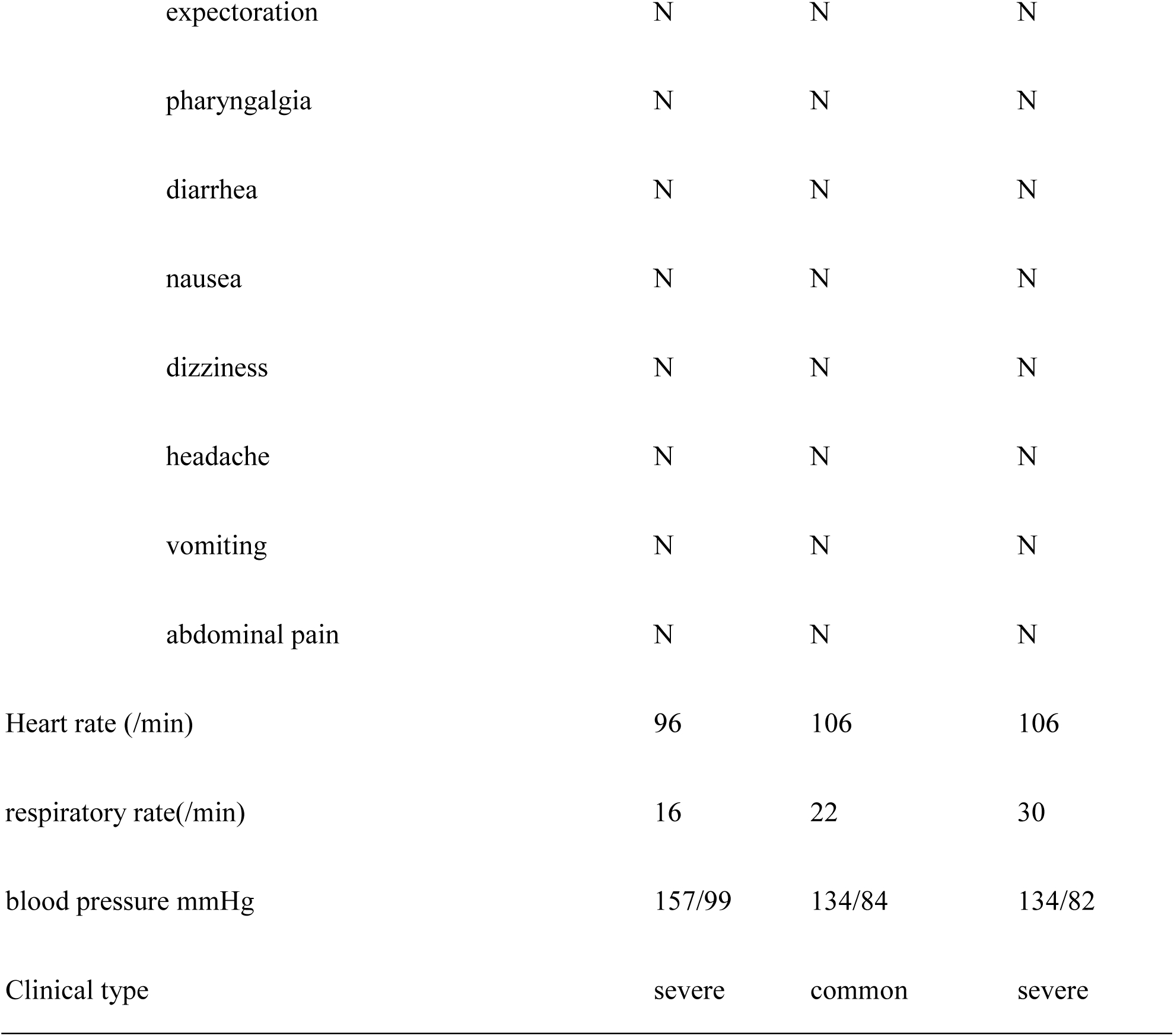
Baseline characteristics of 3 patients with COVID-19

The main laboratory findings of COVID-19 patients were normal or slightly low white blood cells and lymphocytes, elevated inflammatory indicators such as interleukin-6, procalcitonin, C-reactive protein, serum amyloid A, erythrocyte sedimentation rate; and normal myocardial markers (Table 2).

**Table2.** Laboratory findings of 3 patients with COVID-19

|  | Normal<br>range | case1 | case2 | case3 |
| --- | --- | --- | --- | --- |
| white blood cell count,x10<sup>9</sup>/L | 3.5-9.5 | 5.0 | 6.3 | 4.0 |
| neutrophil count,x10<sup>9</sup>/L | 1.9-7.2 | 3.7 | 4.1 | 3.4 |
| Lymphocyte count,x10<sup>9</sup>/L | 1.1-2.7 | 0.9 | 1.7 | 0.4 |
| PaO<sub>2</sub>/FiO<sub>2</sub> mmHg | >300 | 257* | 344 | 110* |
| Aspartate aminotransferase,U/L | 5-34 | 30 | 45* | 53 |
| Alanine aminotransferase,U/L | 0-40 | 22 | 28 | 47* |
| Creatine kinase,U/L | 29-200 | 60.5 | 354.7* | 183.4 |
| Creatine kinase MB isoenzyme,U/L | 0-24 | 16.2 | 16.8 | 36.5* |
| Myoglobin, µg/L | 0-105.7 | 25.5 | 46.9 | 47.8 |
| Troponin I, µg/L | 0-0.04 | 0.00 | 0.01 | 0.01 |
| Interleukin 6,pg/mL | 0-7 | 111.20* | 55.32* | 14.77* |
| Procalcitonin,ng/mL | <0.05 | 0.20* | 0.12* | 0.17* |
| Serum amyloid A, mg/L | <6.8 | 149* | 67* | 217* |
| C-reactive protein,mg/L | 0.0-8.0 | 29.3* | 27.6* | 129.7* |
| D-dimer,ug/L(DDU) | 0-252 | 149 | 213 | 5561* |
| Erythrocyte sedimentation rate,mm/h | 0-15 | 39* | 58* | 73* |
| Creatinine, µmol/L | 59-104 | 79.1 | 106.2* | 63.0 |

\* out of the upper or lower limits

The production of anti-2019-nCoV antibodies after the onset of morbidity is shown in figure 1. Three COVID-19 patients were becoming reactive (positive) for specific anti-2019-nCoV antibodies from 7-12 days after the onset of morbidity, and the levels of anti-2019-nCoV IgM and IgG antibodies increased with the progression of the disease. Closed followed with the production of anti-2019-nCoV IgM antibody, the production of anti-2019-nCoV IgG antibody is also rapid. Of the 3 cases, 1 case developed anti-2019-nCoV IgG 1 day later than IgM, and the other 2 cases developed anti-2019-nCoV IgM and IgG almost on the same day. However, in different cases, the changes of anti-2019-nCoV IgM and IgG were not completely consistent, case 2 showed that the level of anti-2019-nCoV IgG was continue higher than that of IgM, and the other two cases showed that the level of anti-2019-nCoV IgM increased more than that of IgG from 2 weeks of morbidity (Table 3).

**Table 3.**
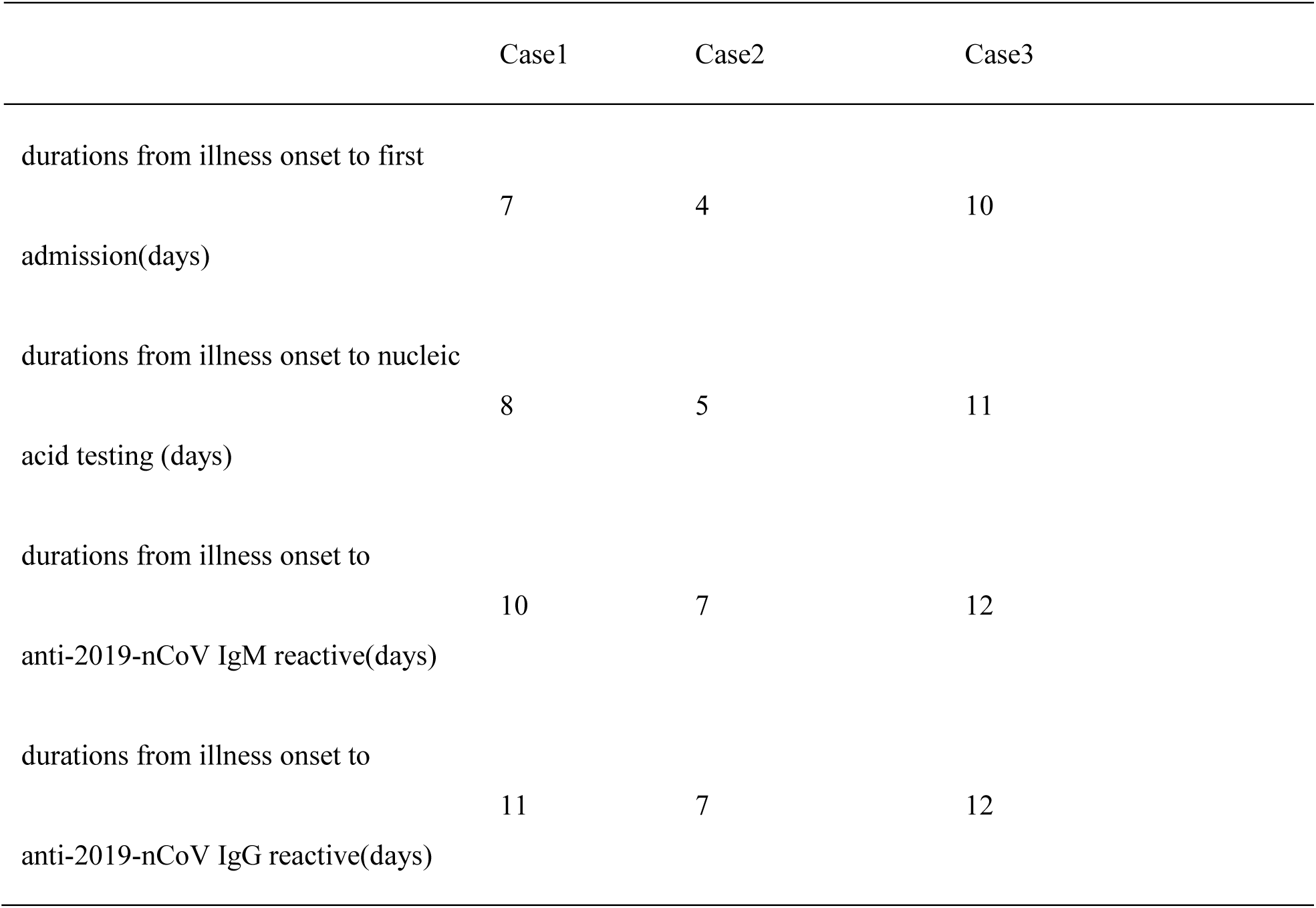
Anti-2019-nCoV antibody production

|  | Case1 | Case2 | Case3 |
| --- | --- | --- | --- |
| durations from illness onset to first admission(days) | 7 | 4 | 10 |
| durations from illness onset to nucleic acid testing (days) | 8 | 5 | 11 |
| durations from illness onset to anti-2019-nCoV IgM reactive(days) | 10 | 7 | 12 |
| durations from illness onset to anti-2019-nCoV IgG reactive(days) | 11 | 7 | 12 |

**Figure 1.**
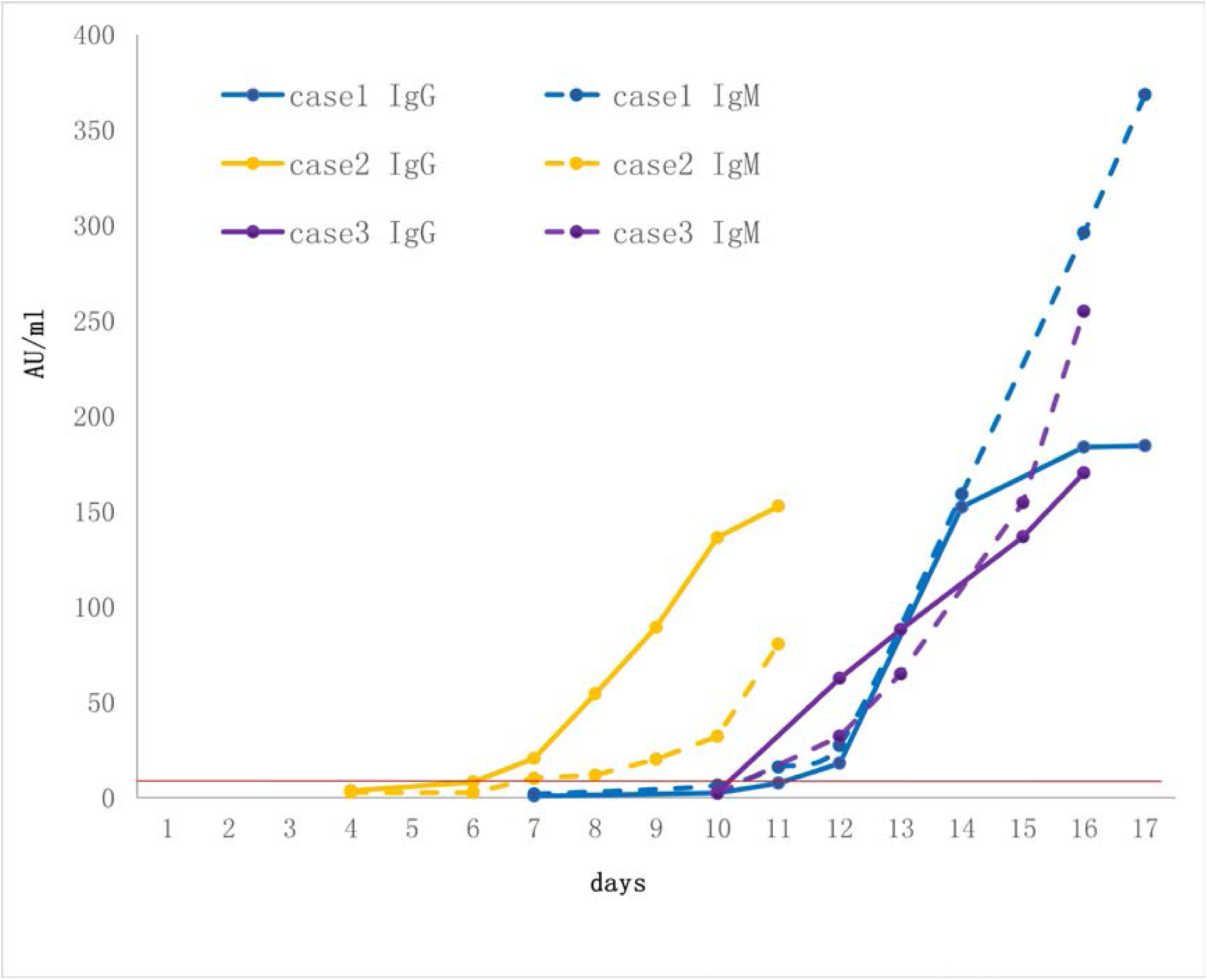
Dynamics of antibody production in 3 patients. The red line represents the positive threshold value, 10.0 AU/ml; The x-axis represents the days since the disease morbidity.

In non-COVID-19, other disease, medical staff and health control groups, there were a few cases reactive for 2019-nCoV IgM and IgG, all the cases were single reactive for IgM or IgG. The sensitivities of IgM and IgG were 100%, as for specificities of IgM and IgG were all over 97% (Table 4).

**Table4.** Anti-2019-nCoV antibody detection in different groups

|  | non-COVID-19 | other Disease | medical staff | health control |
| --- | --- | --- | --- | --- |
| number | 225 | 222 | 63 | 223 |
| age years,median(range) | 35(1-86) | 50(27-85) | 40(25-61) | 59(21-95) |
| male/female | 124/101 | 62/160 | 7/56 | 77/146 |
| 2019-nCoV IgM reactive | 6 | 2 | 0 | 3 |
| 2019-nCoV IgG reactive | 1 | 2 | 0 | 4 |
| 2019-nCoV IgM median/P99(AU/ml) | 1.82/19.66* | 0.85/10.99 | 1.37/4.56* | 0.86/11.35 |
| 2019-nCoV IgG median/P99 (AU/ml) | 1.82/8.52 | 1.21/10.52* | 1.27/6.26 | 1.49/11.18 |
| 2019-nCoV RNA | 0 | N/A | N/A | N/A |
| influenza A RNA | 2 | N/A | N/A | N/A |
| influenza B RNA | 2 | N/A | N/A | N/A |
| adenovirus DNA | 4 | N/A | N/A | N/A |
| mycoplasma pneumoniae DNA | 17 | N/A | N/A | N/A |
| Sensitivity(IgM) | 100% | 100% | 100% | 100% |
| Sensitivity(IgG) | 100% | 100% | 100% | 100% |
| specificity(IgM) | 97.33% | 99.10% | 100.00% | 98.65% |
| specificity(IgG) | 99.56% | 99.10% | 100.00% | 98.21% |
| negative predictive values(IgM) | 100.00% | 100.00% | 100.00% | 100.00% |
| positive predictive values(IgM) | 33.33% | 60.00% | 100.00% | 50.00% |
| negative predictive values(IgG) | 100.00% | 100.00% | 100.00% | 100.00% |
| positive predictive values(IgG) | 75.00% | 60.00% | 100.00% | 42.86% |

\*compared with health control, $P<0.05$

Of 225 non-COVID-19 cases, 2 cases were detectable for influenza A RNA and 2 cases were detectable for influenza B RNA, respectively, 4 cases were detectable for adenovirus DNA, 17 cases were detectable for mycoplasma pneumonia DNA (Table 4).

We also compared the anti-2019-nCoV antibodies values distributions in different groups. The anti-2019-nCoV IgM levels in non-COVID-19 was higher than that of healthy control group, the difference was statistically significant (Table 4; Figure 2).

**Figure 2.**
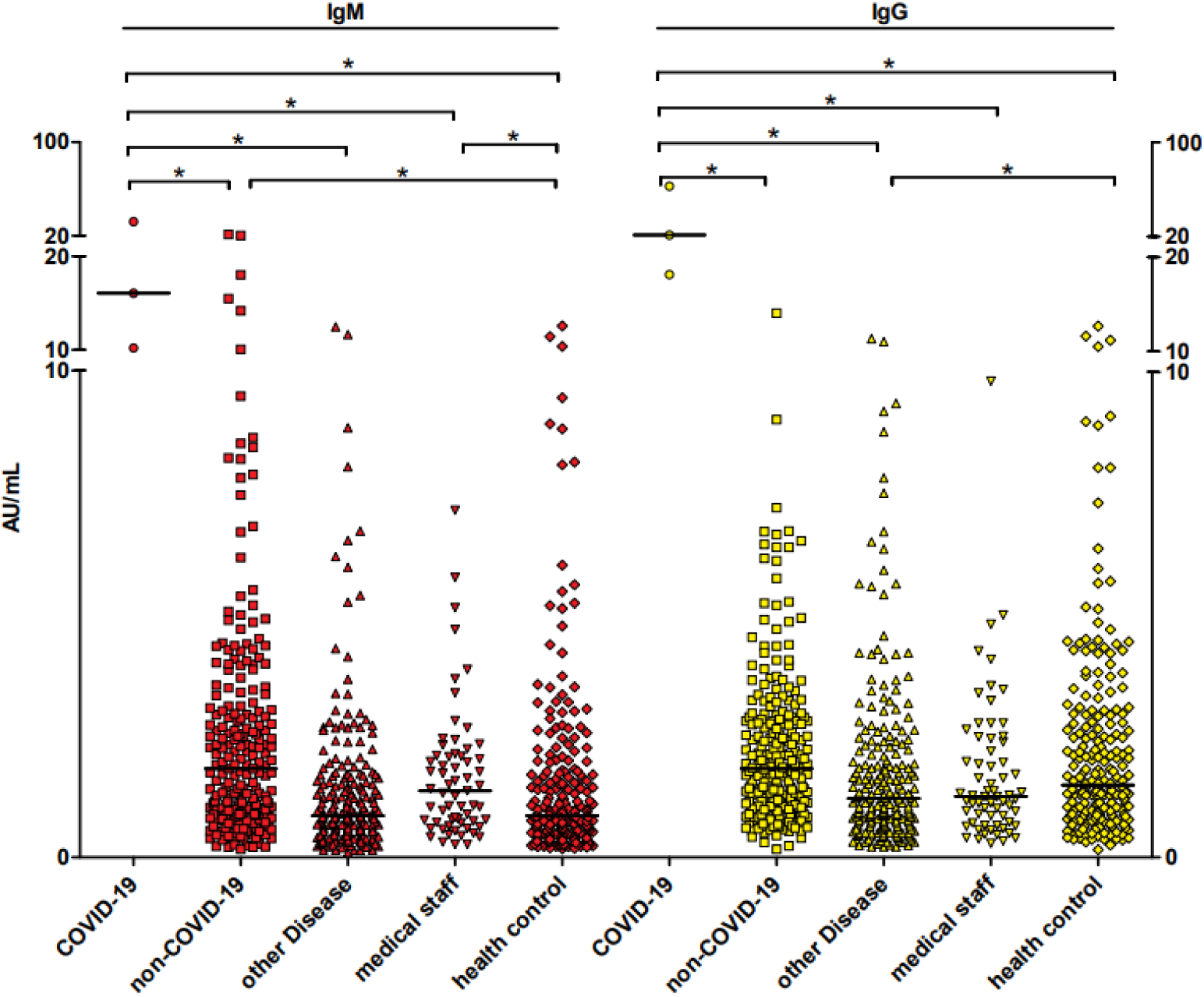
The anti-2019-nCoV IgM and IgG antibodies distribution in different groups. Each data point represents the antibody level of the participants, the short horizontal line represents the median antibody level of the group, and * represents the difference between the two groups is statistically significant, *P*<0.05.

In order to further clarify the diagnostic efficacy of specific IgM and IgG in fever suspected COVID-19 patients (all 228 patients had been proved by testing 2019-nCoV nucleic acid), we made the ROC curve of anti-2019-nCoV IgM and IgG. The area under the curve was 0.988 and 1.000, and the best cut-off value was 10.14 and 15.99, respectively (Figure 3).

**Figure 3.**
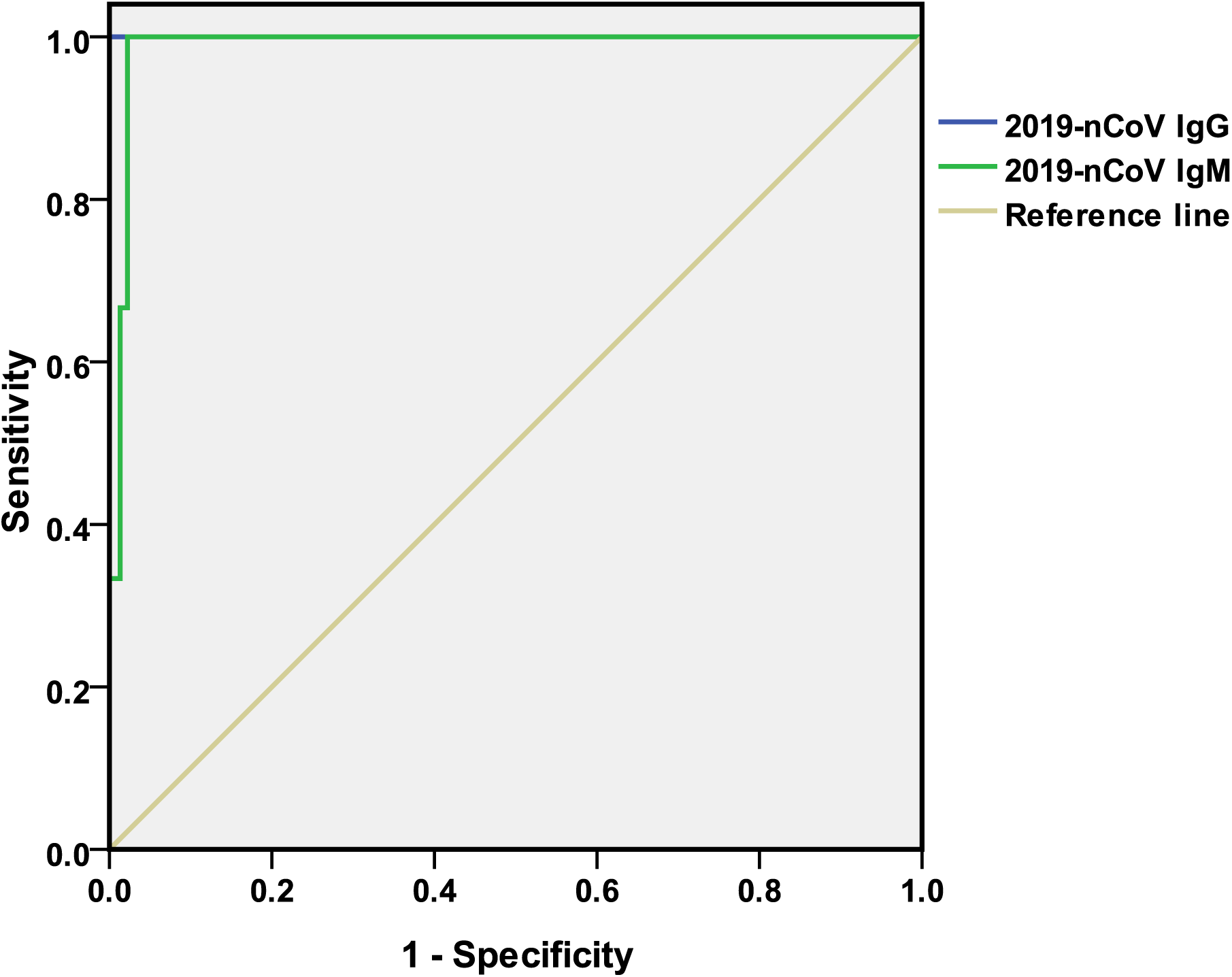
ROC curves of anti-2019-nCoV IgM and IgG in the diagnosis of COVID-19.

## Discussion

With China’s growing surveillance network and laboratory capacity, the outbreak was identified within a few weeks and the viral genome sequence was announced (*10*), effectively promoting in vitro diagnostic tests. At present, the main diagnostic method is to detect 2019-nCoV nucleic acid by real-time quantitative fluorescent PCR.

In the early stage of this epidemic, due to the insufficient production of nucleic acid detection kits and the high requirement of technical norms for nucleic acid detection, the application of nucleic acid testing as a diagnostic standard was limited, and not all suspected patients could be tested in time for definite diagnosis. With the efforts of China State Drug Administration, seven 2019-nCoV nucleic acid detection reagents have been urgent approved to be marketed. It effectively alleviated the problem that the lack of reagents in the epidemic prevention and control. At mean time, with the number of COVID-19 cases increased, physicians found that only 30-50% of confirmed cases tested positive for 2019-nCoV nucleic acid, which means there was a large part of patient had missing detection of nucleic acid. The factors owing to this missing include the timing of oropharyngeal or nasopharyngeal specimens’ collection; improper collection site, such as the collection depth was not enough; the urgent launch of a large number of in vitro diagnostic reagents inevitably leads to the lack of large sample virus genome research and clinical validation; standardized clinical nucleic acid testing laboratory was still a limitation to all labs. Furthermore, in the course of COVID-19’s different stages, the virus load is different. When the body’s immune system produces antibodies, the clinical signs and symptoms is obvious, the virus is likely to decline without being detected.

Recently, a study of 138 patients showed a high proportion (41%) of suspected nosocomial infection (*11*), and it is also noteworthy that the presence of asymptomatic patients with latent mild pneumonia may be an important source of infection for outbreak transmission (*12*). Therefore, it remains critical to apply a fast and convenient detection method to distinguish and trace suspicious case or contacts as early as possible in order to prevent super-transmission events.

Antibodies are the products of humoral immune response after infection with viruses. As a new infectious disease, while the detection of nucleic acid cannot be used widely, specific antibodies to 2019-nCoV can be used to determine whether the patient has been recently infected with 2019-nCoV or not. It has been reported that serum samples from 5 patients were detected by self-made 2019-nCoV IgG and IgM ELISA kits, and the antigen could cover 92% of the 2019-nCoV NP amino acids (*13*).

Generally speaking, the immune response of pathogenic microorganisms is usually stimulated by the rise of IgM after infection, IgG usually appears 1-2 weeks after IgM, and has been rising and maintaining high levels in the body for a long time. Because COVID-19 is a new infectious disease and the immunological test reagent has just been developed, there is still no report on how IgM and IgG antibodies were produced and developed after 2019-nCoV infection.

In our study, we found that specific antibodies reactive to 2019-nCoV appeared from 7-12days after the onset of morbidity in all 3 patients. Unlike previous experience that IgG usually appears 1-2 weeks after IgM, the presence of anti-2019-nCoV IgM antibodies in COVID-19 cases was followed by the presence of anti-2019-nCoV IgG antibodies within a very short period of time (about 0∼1 day), followed by a simultaneous rise in both antibodies. However, the rising speeds of anti-2019-nCoV IgG and IgM antibodies were different in different individuals. It was very interesting to find that the case2, who had the mildest clinical signs and symptoms among the 3 COVID-19 patients, was the earliest patient to show the specific anti-2019-nCoV antibodies on 7^th^ day of morbidity, his anti-2019-nCoV IgM value was relative lower but IgG value was relative higher in the following days; on the other hand, Case 3, who had the severest clinical signs and symptoms among the 3 COVID-19 patients, was the latest patient to show the reactivity to anti-2019-nCoV on the 12^th^ day of morbidity, and the value of anti-2019-nCoV IgM antibodies continued to increase.

2019-nCoV is highly infectious and the population is generally susceptible to 2019-nCoV.The most common symptoms after infection include fever, fatigue, dry cough, and muscle pain, with expiratory dyspnoea occurring in more than half of patients (*14*). Severe cases are prone to rapidly progress to acute respiratory distress syndrome, septic shock, high risk of admission to intensive care units, and even death. Therefore, how to closely observe the condition after morbidity and find severe cases as soon as possible is the key to reduce the mortality of critically ill patients. According to our findings, it seems that the time and speed of production of specific anti-2019-nCoV IgM antibodies correlate with disease severity. But because the number of cases is so small, more research is needed to confirm it.

In addition to COVID-19, the fever patient of non-COVID-19, other disease, medical staffs and healthy controls were also studied. Non-COVID-19 group included several other respiratory viruses such as influenza A, B and adenovirus infection cases, these cases were negative for anti-2019-nCoV specific antibody detection, indicating that the antibody detection has a good ability to resist interference and differential diagnosis of different respiratory virus infections. Among each groups, only COVID-19 patients were all positive for both anti-2019-nCoV IgM and IgG antibodies, while in other populations, the IgG or IgM antibodies were single positive in a very few cases. However, combined with the results of 2019-nCoV nucleic acid detection and clinical data, they were judged to be a false positive of IgM or IgG. Considering that COVID-19 has broken out in many countries around the world, more than 80000 people have been diagnosed and the number is growing rapidly, the main problem at present is the need for highly sensitive tests to screen the suspected cases and to prevent missed diagnosis by nucleic acid tests, lower false positive rates for antibody testing are acceptable. In the meantime, for patients with morbidity for a week or more, simultaneous positive of anti-2019-nCoV IgM and IgG will be helpful to improve the specificity.

Compared with nucleic acid test, which requires respiratory tract samples and complex testing procedures, the operation requirement of serum antibody detection in clinical laboratory is lower than that of nucleic acid detection, which can be detected quickly (30min) and in large quantities, and can be completed in common P2 Biosafety Laboratory. When the morbidity is more than a week, nucleic acid detection is not convenient, serological dynamic monitoring can be carried out, once positive, it is strongly recommended to use nucleic acid diagnosis immediately.

The disadvantage of nucleic acid detection is the existence of relative high false negative rate, and serological antibody detection has the advantage of high sensitivity, so the combination of the two will be a good diagnostic means. It can be inferred that after the future epidemic situation has been controlled to a certain extent, as a convenient method, antibody detection is still necessary to make differential diagnosis of other respiratory pathogens infection.

It must be emphasized that independent results of specific antibodies testing should not be used as a diagnostic criteria, especially when the epidemiological history is unclear, and must be combined with the patient’s morbidity time and clinical signs.It must be emphasized that independent results of specific antibodies testing should not be used as a diagnostic basis, especially when the epidemiological history is unclear, and must be combined with the patient’s morbidity time and clinical signs.

To our knowledge, little has been reported about the specific antibody production process in the course of COVID-19 disease, and little has been reported about the different situation of antibodies in fever non-COVID-19 population, other diseases, special contact population such as medical staff and healthy population. This study provides data on the regularity of antibody production in the course of COVID-19, and provides some understanding of the basic data of specific antibodies in different populations. The results of this study help to provide evidence for rapid screening of suspected cases through the serological testing to curb the rapid progress of the epidemic globally. Just on the day of this manuscript was submitted, the China National Health Commission published the new edition of “Diagnosis and Treatment plan of Corona Virus Disease 2019” (*15*), in which recommends that positive of anti-2019-nCoV IgM and IgG can be used as diagnostic criteria, supported our findings.

This study still has some limitations. First, only 3 confirmed COVID-19 cases were included, and although the continuous dynamic process of anti-2019-nCoV antibody production and its relationship with disease progression have been carefully observed, a large sample of cases is still needed for verification. Second, changes in anti-2019-nCoV antibodies were only tracked for 4 to 20 days after the morbidity, with no longer-term observation. However, the trend of anti-2019-nCoV IgG and IgM antibodies production from beginning to increasing has been preliminarily found. Third, anti-2019-nCoV nucleic acid testing has not all been performed in every groups, asymptomatic infections may be missed in other disease group, which might have a certain impact on the evaluation of the diagnostic efficacy of antibodies. Considering that Liaoning Province, where this study was conducted, is a low epidemic area, the possibility of asymptomatic infection would be very small.

In this paper, we studied the producing process of specific antibody in patients with COVID-19, compared and evaluated the diagnostic value of antibody in different populations, which is beneficial for doctors to use in the process of diagnosis and treatment. As a useful complement to nucleic acid detection, the detection of specific anti-2019-nCoV antibodies will be able to draw a more comprehensive, rapid and accurate diagnosis to COVID-19, so as to effectively distinct between COVID and non-COVID-19 patients and curb the rapid spread of 2019-nCoV in the global epidemic period.

## Data Availability

The data used to support the findings of this study are available from the corresponding author upon request.

